# Impact of Radiation Safety Training and Years of Experience after Cardiology Training on Radiation Safety Practices : An Alarming Trend identified and Solutions offered!

**DOI:** 10.64898/2025.12.01.25341422

**Authors:** Gnanaraj Justin Paul, Ashok Kumar, Anne Princy Steaphen, Dhamodaran Kaliyamoorthy, Dorairaj Prabhakar, Harilalith Kovvuri, Janakiraman Ezhilan, Kesavamoorthy Boopalan, Kumaresan Kannan, Marimuthu S Ravi, Prathap Kumar N, Pratap Kumar Gorijavaram, Sadanand Shetty, Sundar Chidambaram, Swethrajan Gunasekaran, Winfred Gnanaraj Justin Paul, Nagendra Boopathy Senguttuvan

## Abstract

**Background:** Occupational radiation exposure remains a persistent and underestimated hazard in cardiac catheterization laboratories. We evaluated the impact of post-licensure radiation safety training on adherence to radiation safety practices among interventional cardiologists

**Methods:** A nationwide cross-sectional survey was conducted between May 2022 and August 2023 among 753 interventional cardiologists Radiation safety knowledge and practise were assesses across five domains - dose awareness, time, distance, shielding, and collimation - and combined into a composite Radiation Safety Score (RSS), with a maximum possible score of 100. Demographic and practice-related variables were analyzed, and multivariable log-linear regression was used to identify independent predictors of higher RSS.

**Results:** Of the 753 respondents, 716 met inclusion criteria. The median age was 44 years (IQR 37–52), and the mean catheterization laboratory experience was 11 ± 8.7 years. Two-thirds (66%) had attended at least one post-licensure radiation safety training program. The mean RSS was 56.5 ± 21.7, and only 16.3% achieved scores ≥ 75. Trained participants demonstrated significantly higher performance across all five radiation safety domains (p < 0.001 for all comparisons). In multivariable analysis, post-licensure radiation safety training independently predicted higher RSS (β = 0.216, p = 0.005), corresponding to a 24.1% higher adjusted score. Experience >10 years was also an independent predictor of superior RSS (β = 0.267, p < 0.001).

**Conclusion:** Radiation safety practices among interventional cardiologists in India are suboptimal, and requires substantial improvement. Participation in post-licensure radiation safety is independently associated with significantly better adherence across all major radiation safety domains. Incorporating structured radiation safety lectures and workshops into cardiology conferences- particularly targeted at early-career interventional cardiologists —may help strengthen safety culture and improve long-term occupational protection.

## Introduction

Advances in interventional cardiology have transformed the management of cardiovascular disease by enabling minimally invasive treatment for coronary artery disease, structural heart defects, and electrophysiological abnormalities, to name a few. However, these procedures rely heavily on fluoroscopy and X-ray imaging, exposing both patients and personnel to ionizing radiation. Unlike patients, for whom exposure is episodic, healthcare professionals in the catheterization laboratory sustain cumulative, low-dose radiation exposure over a career spanning decades, which confers upon them a significantly elevated lifetime risk of stochastic effects such as malignancy, as well as deterministic injuries such as cataracts, skin damage, and musculoskeletal disorders associated with prolonged adornment of protective gear.^1–3^ In response to these direct and indirect hazards of radiation, the "As Low as Reasonably Achievable" (ALARA) principle was established to operationalize fundamental safety practices by emphasizing awareness of radiation time, distance, shielding, and dose. Despite the existence of established international guidelines, adherence to best radiation safety practices in catheterization laboratories remains suboptimal worldwide. For instance, a recent cross-sectional study at a tertiary care center in India found that 87% of cardiac catheterization (cath)lab staff had no formal radiation safety training, and less than half correctly identified the annual occupational dose limit or understood basic principles such as the inverse square law.^4^ Remarkably, 56% of the staff admitted to forgoing personal protective equipment altogether for reasons related to perceived discomfort or “inconvenience”. These results are broadly consistent with trends from around the world, which have highlighted deficiencies among cath lab personnel in awareness of institutional dose monitoring protocols and individual exposure limits, usage of additional protective equipment such as goggles and personal dosimeters, and in participation rates of radioprotection programs.^5,6^ This gap suggests that foundational knowledge acquired during training is insufficient to instil durable safety behaviours. In India, while some radiation safety is taught during education, (through textbooks and occasional lectures and didactic sessions) such training is not uniform, nor is it mandatory in cardiology curricula. Consequently, many early-career interventionalists enter the laboratory with skills gap, focusing predominantly on mastering complex techniques while potentially underestimating the long-term occupational consequences of poor radiation safety practices. Post-licensing initiatives present a valuable opportunity to address this knowledge-to-practice gap. Such targeted training can reinforce core principles, emphasize dose-reduction practices, and cultivate a culture of safety for cath lab professionals and patients. To this end, we have delivered structured radiation safety training lectures at major interventional cardiology conferences across India, wherein our educational approach emphasized practical and feasible implementation strategies that were distilled into "Ten Commandments of Radiation Safety”. These commandments focused on five fundamental concepts: dose awareness, time optimization, distance maximization, appropriate shielding, and collimation. The present study investigated the impact of these post-licensing radiation safety training lectures by administering a knowledge, attitudes, and practices (KAP) survey to interventional cardiologists across India and aimed to assess the relationship between structured radiation safety training and adherence to evidence-based protection practices.

## Methods

### Study Design and Population

We conducted a prospective, cross-sectional survey of interventional cardiology professionals across India between May 2022 and August 2023. The study was approved by the Institutional Ethics Committee at Madras Medical College (Ref No: 01102022) and written informed consent was obtained from all the participants.

### Survey Instrument and Data Collection

A questionnaire was pilot-tested among a small sample of interventional cardiology practitioners and refined for clarity and validity prior to deployment. The survey was hosted on Google Forms and distributed to participants via two channels. The survey was directly administered at two national and one regional cardiology conferences, and was also disseminated electronically via professional social media groups comprising of interventional cardiologists. Duplicate/repeat responses from the same participant were identified and excluded from the analysis. The survey incorporated multiple-choice and Likert-scale questions designed to assess knowledge and practices related to five fundamental elements of radiation protection as defined by international standards. 1) Questions addressing time evaluated interventionalists’ knowledge and implementation of strategies to minimize exposure duration and intensity. 2) Distance questions assessed understanding and application of optimal positioning techniques for operators, patient tables, and detectors to maximize radiation safety according to the inverse square law principle. 3) Shielding questions evaluated the effective use of available protective equipment to safeguard operators and staff from scatter radiation. 4) Collimation questions focused on proper use of collimation techniques to minimize primary radiation exposure to the patient. 5) Dose awareness questions assessed practices related to monitoring, recording, and reporting radiation exposure levels as mandated by institutional and/or international safety recommendations. The survey also collected basic demographic details and professional characteristics. The latter included total years of experience in the catheterization laboratory, field of sub-specialization, self-reported illnesses or conditions attributed to radiation exposure, and history of any post-licensure radiation safety training (which was defined as lectures, workshops, and/or formal courses attended after qualification).

### Radiation Safety Score Development

To quantify overall radiation safety performance, we developed a composite Radiation Safety Score (RSS) with a maximum of 100 points. Each of the five radiation safety domains (time, distance, shielding, collimation, and dose awareness) was assigned an equal weight of 20 points.

### Outcome Measures

The primary outcome was the impact of post-licensure radiation safety training attendance on radiation safety practices, as measured by the comprehensive Radiation Safety Score. The secondary outcome evaluated the relationship between years of experience in cardiac catheterization laboratories and radiation safety practice adherence.

### Statistical Analysis

The responses of all participants were included in the primary analysis, while responses from nurse practitioners and cardiac catheterization technicians were excluded from the comparative analysis. Continuous variables are presented as mean ± standard deviation or median (interquartile range [IQR]) based on normality of the data, as assessed by the Shapiro-Wilk test. Between-group comparisons of continuous variables were performed using independent t-tests for normally distributed data or Mann-Whitney U tests for non-parametric data. Categorical variables are expressed as frequencies and percentages and were analyzed using Chi-square or Fisher’s exact test. A multivariable log-linear regression model was developed to assess the independent impact of a few key variables on the overall Radiation Safety Score while controlling for potential confounding variables. Selected covariates included sex, prior radiation safety training exposure, years of experience in interventional cardiology, employment sector (public vs private hospital), as well working in a teaching versus non-teaching hospital. All statistical analyses were performed using SPSS version 23.0 (IBM Corporation, Armonk, NY, USA) and Stata version 18 (StataCorp LLC, College Station, TX, USA). A two-sided p-value <0.05 was considered statistically significant for all variables of interest and was reported alongside 95% confidence internals (CIs).

## Results

### Baseline Clinical Characteristics and Radiation Safety Practices

A total of 753 interventional cardiologists across India participated in the survey. After excluding responses from 37 participants who did not meet inclusion criteria or provided incomplete data, 716 respondents were included in the final analysis. Of these 95% (682) were adult interventional cardiologists, while paediatric interventional cardiologists and cardiac electrophysiologists constituted 2% (13) and 3% (21), respectively. The cohort was predominantly male (88.8%) with a median age of 44 years [IQR 37-52]. The median duration of professional experience in the catheterisation laboratory was 10 years [IQR 5-17], with comparable representation across experience levels. 22.3% (160) of respondents had less than 5 years of experience, while 33.2%(238) had between 5-10 years, and 44.4% (318) had more than 10 years of experience. Nearly two-thirds of participants (n=472, 65.9%) reported attending at least one formal radiation safety lecture, workshop, or training program after obtaining their cardiology licensure. Attendance was significantly associated with years of experience and practitioners with >10 years of experience were more likely to have attended such training compared to those with ≤10 years of experience (74.8% versus 58.8%; p < 0.001). Regarding the use of shielding measures, thyroid collar was the most consistently used personal shielding device (84.2%), followed by the under-table shield (63.3%) and ceiling-mounted suspended shield (58.1%) **(Figure 1)**.

**Figure 1.**
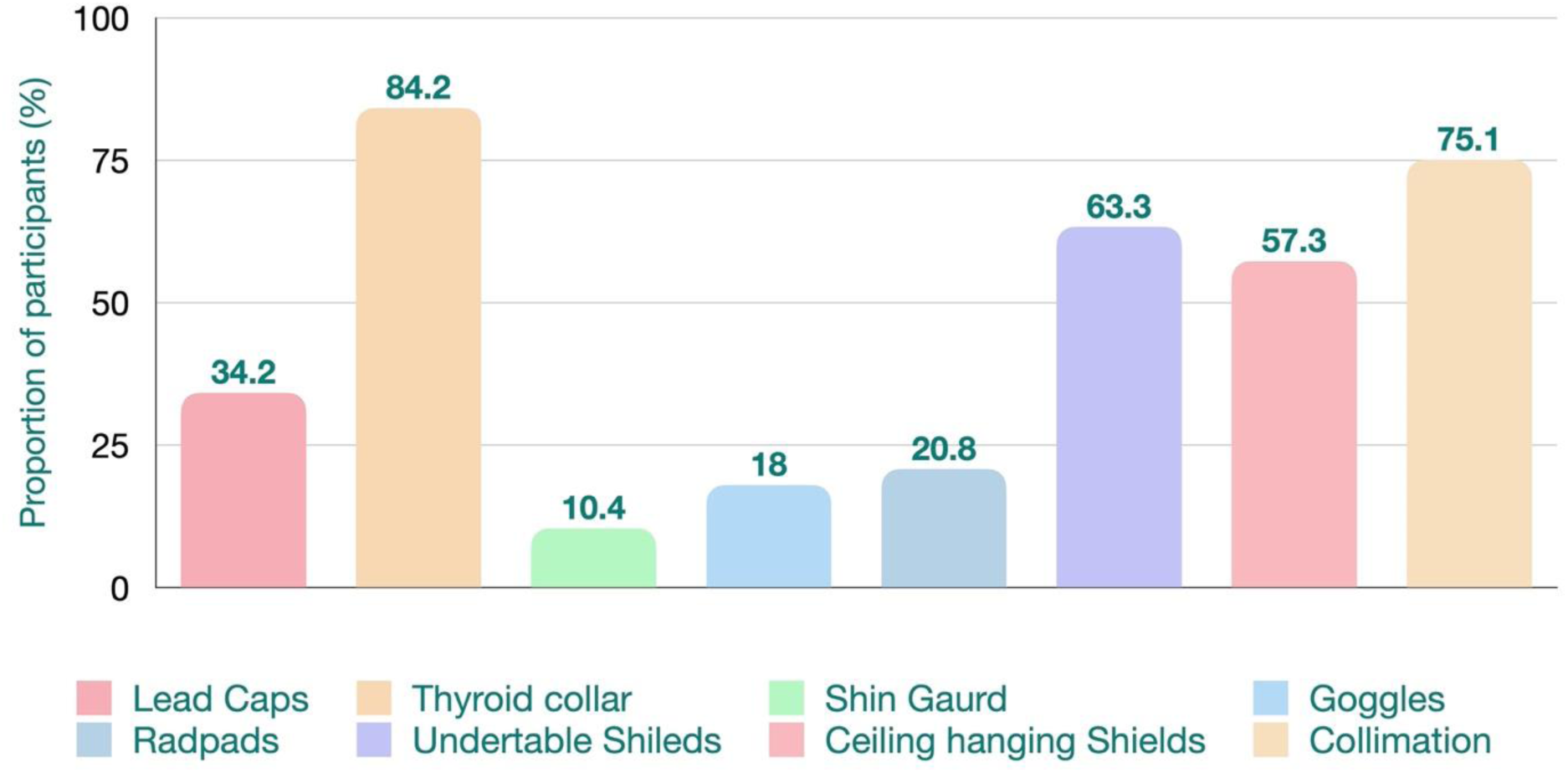
Use of radiation protection by the study participants

Collimation during procedures was practiced by approximately three-fourths of respondents (446). About 58.4% reported using fluoroscopy storage features like Fluoro Store or Fluoro Save, while only 45.3% reported using frame rates below 15 frames per second. Personal radiation dose monitoring was reported by two-thirds of respondents, however, less than one-quarter documented radiation doses in procedural reports. When asked about preferences for post-licensing training programs, 68% of participants recommended lectures be conducted for all attendees at interventional cardiology conferences across the nation. An additional 20% favoured inclusion of online certification courses following the training program, while 11% felt that such lectures were necessary only during dedicated fellows’ courses at conferences and not for all attendees.

### Effect of Radiation Safety Training on Safety Scores

The mean composite RSS for the entire cohort was 56.46 ± 21.66 out of a maximum of 100 points. Only 117 participants (16.3%) achieved a score ≥75, while 385 (53.7%) registered scores between 50-74. A concerning 214 interventionists (30%) scored below 50, indicating that a substantial proportion of participants had suboptimal radiation safety competency. Consistent with our hypothesis, post-licensure radiation safety training had a significant positive impact on radiation safety practices. Participants who had attended at least one radiation safety training program (n=472) achieved significantly higher median RSS (63.6 [IQR 52.7-74.7]) compared to those without any post-licensure training (52.2 [IQR: 37.8-67.2]; p < 0.001). Among the participants with radiation safety training, 77.5% achieved RSS ≥50, compared to only 54.3% of untrained participants (p < 0.001). Domain-specific analysis revealed that the benefit of post-licensure training was consistent across all five fundamental radiation safety principles, with trained participants demonstrating superior performance in every domain (p < 0.001 for all comparisons) **(Figure 2).**

**Figure 2.**
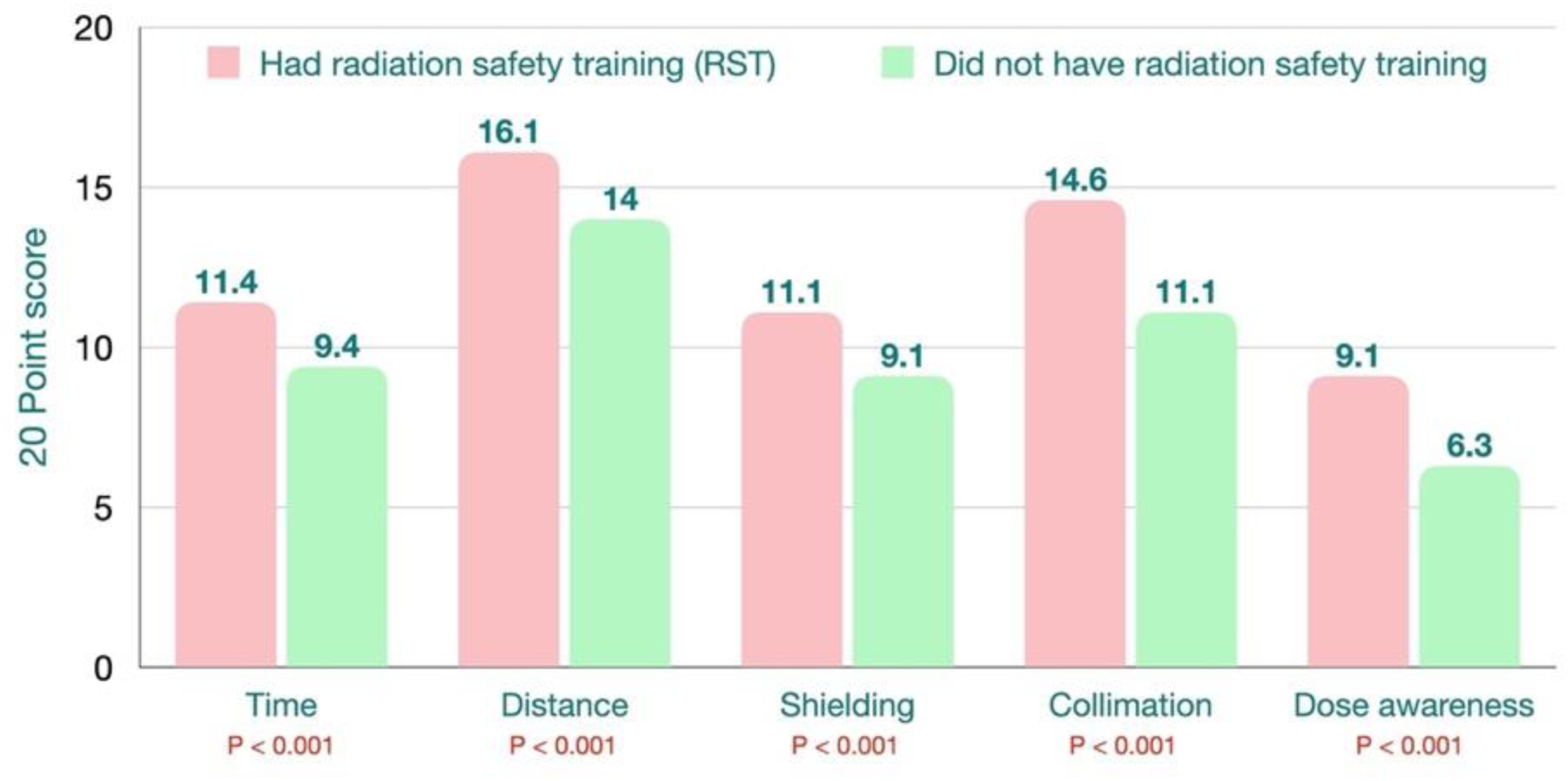
Influence of post licensure RST on five key concepts of RSP

### Effect of Radiation Safety Training on Individual Radiation Safety Practices

Participants who attended radiation safety training demonstrated significantly better adherence to key radiation safety practices compared to those without such training (Table-1). The use of dosimeters was markedly higher in the trained group (72.4% vs. 50.4%; p < 0.001), as was personal radiation dose monitoring (75.4% vs. 48.4%; p < 0.001) and documentation of radiation doses in procedure reports (25.4% vs. 19.3%; p < 0.001). Use of protective equipment followed the same pattern. Trained participants more frequently used thyroid collars (86.7% vs. 79.5%, p=0.013), ceiling-mounted suspended shields (62.3% vs. 46.3%, p<0.001), under-table shields (68.4% vs. 53.3%, p<0.001), lead goggles (20.3% vs. 13.5%, p=0.025), and disposable radiation-absorbing pads (RadPads) (23.7% vs. 15.2%, p=0.007). Collimation, routine calibration of angiographic equipment at 6 to 12-month intervals, positioning the image intensifier closer to the patient, and consciously standing away from the X-ray tube during exposure were also more frequently reported to be performed by those who attended at least one training session (p < 0.001 for all comparisons). Although trained operators more frequently used fluoroscopy storage features like Fluoro Store or Fluoro Save (62.9% vs 49.6%, p < 0.001), use of lower frame rates (≤ 7.5 frames per second) did not differ significantly between the groups (46.6% vs 42.6%, p=0.31). Taken together, these results indicate that post licensure safety training is associated with higher uptake of both fundamental and more technical radiation-safety measures.

### Effect of Professional Experience on Individual Radiation Safety Practices

Operators with more than ten years of experience in the catheterization laboratory achieved significantly higher radiation safety scores compared to those with less experience (65.6 ± 14 vs. 52.8 ± 24; p < 0.001). Among operators with more than ten years of experience, 79% (260) achieved a radiation safety score > 50, whereas only 62.3% (264) of operators with less than ten years of experience reached this threshold (p < 0.001). More experienced operators performed significantly better across most domains, including time, distance, shielding, collimation and dose awareness domains(Table-2). However, no significant differences were observed between the two groups for specific practices such as use of RadPads, lead goggles, headgear, shin guards, or preference for very low frame-rate fluoroscopy

### Multivariable log-linear regression analysis

Factors associated with higher radiation scores were evaluated using multivariable log-linear regression. Post-licensure radiation safety training was independently and strongly associated with higher RSS (β = 0.216, p = 0.005), corresponding to an estimated 24.1% increase in RSS among trained operators. More than ten years of experience in the cath lab was associated with higher RSS (β=0.267, p < 0.001), reflecting a 30.6% increase, compared to those with less than ten years of experience. Staff at teaching institutions demonstrated significantly better radiation safety adherence than those at non-teaching institutions (β=0.165, p=0.048), representing an estimated 17.9% higher RSS. There was no significant difference in radiation safety practices based on gender. However, practice at public institutions was associated with a positive but non-significant increase in RSS (β = 0.151, p=0.153). No significant differences in RSS were observed based on gender. (Table 3).

**Table 1:**
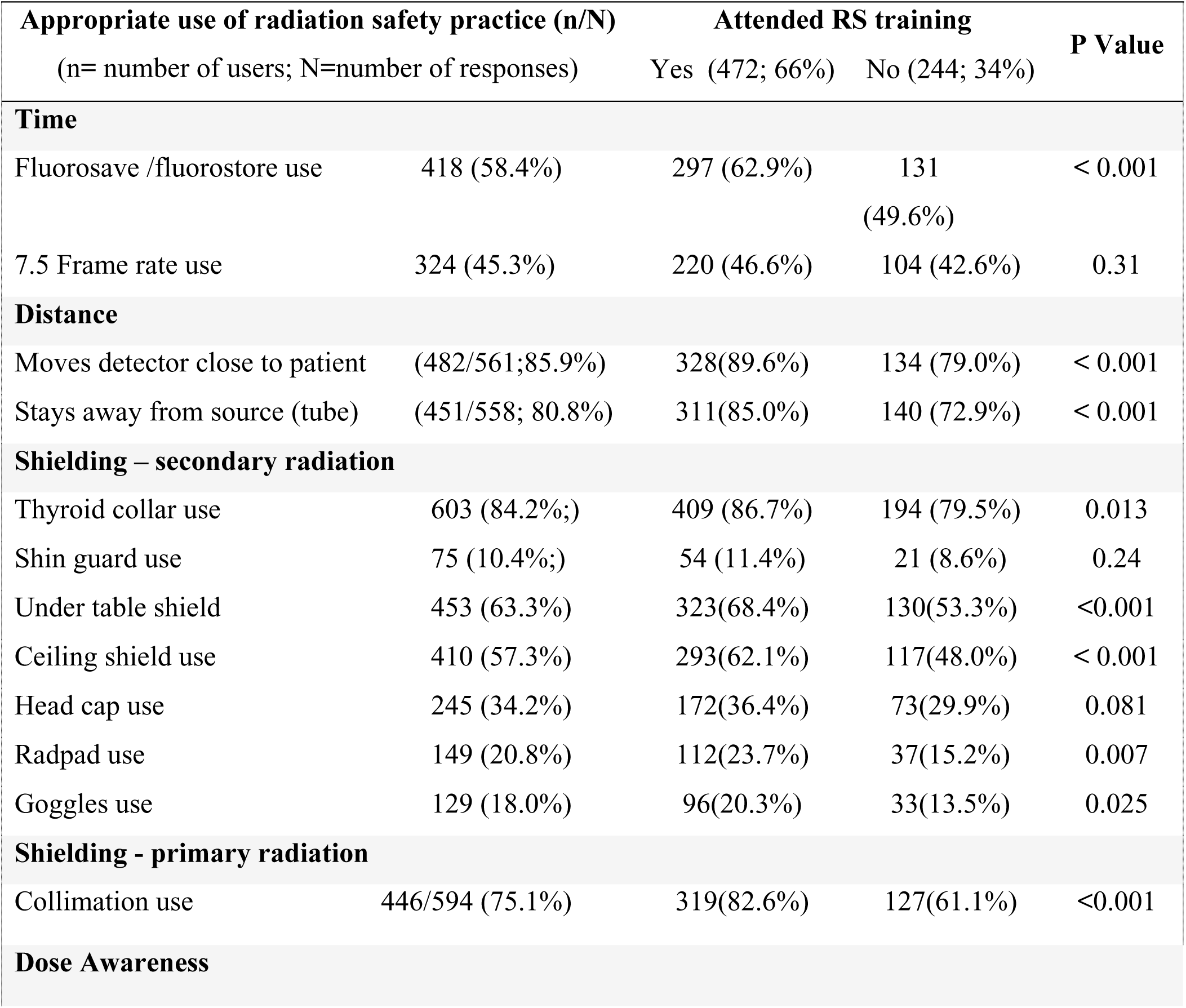
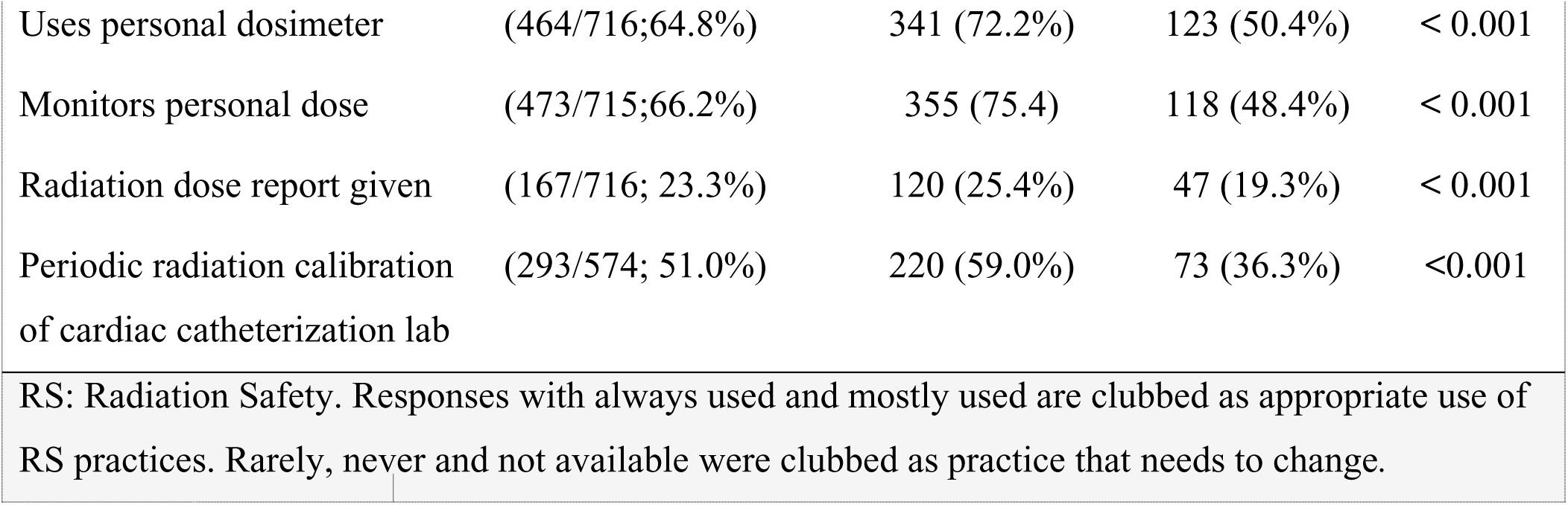
Effect of Radiation safety (RST) training on Radiation safety practices (RSP)

**Table 2:** Effect of years of experience in catheterisation laboratories on RS practices.

| Radiation safety practice (n/N) |  | Years of experience |  | P Value |
| --- | --- | --- | --- | --- |
| (n= number of users; N=number of responses) |  | <10 years<br>(398; 56%) | > 10 years<br>(318; 44%) |  |
| Dose Awareness |  |  |  |  |
| Uses personal dosimeter | 464/716; 64.8% | 193 (48.5%) | 271 (85.2%) | < 0.001 |
| Radiation dose monitoring | 473/715; 66.2% | 214 (53.8%) | 259 (81.7%) | < 0.001 |
| Radiation dose report given | 167/716; 23.3% | 98 (24.6%) | 69 (21.7%) | < 0.001 |
| Cath lab calibrated<br>periodically | 293/574; 51.0% | 152 (43.7%) | 141 (62.4%) | < 0.001 |
| Time |  |  |  |  |
| Fluorosave /fluorostore use | 418/716; 58.4% | 203 (51.0%) | 215 (67.6%) | < 0.001 |
| 7.5 Frame rate use | 324/716; 45.3% | 172 (43.2%) | 152 (47.8%) | 0.221 |
| Distance |  |  |  |  |
| Moves detector close to<br>patient | 482/561; 85.9% | 275 (81.6%) | 207 (92.4%) | < 0.001 |
| Moves away from source | 451/558; 80.8% | 246 (73.9%) | 205 (91.1%) | < 0.001 |

| Shielding from secondary radiation |  |  |  |  |
| --- | --- | --- | --- | --- |
| Thyroid collar use | 603/716; 84.2% | 325 (81.7%) | 278 (87.4%) | 0.036 |
| Under table shield use | 453/716; 63.3% | 225 (56.8%) | 227 (71.4%) | < 0.001 |
| Ceiling shield use | 410/716; 57.3% | 204 (51.3%) | 206 (64.8%) | < 0.001 |
| Head cap use | 245/716; 34.2 | 141 (35.4%) | 104 (32.7%) | 0.465 |
| Radpad use | 149/716; 20.8% | 90 (22.6%) | 59 (18.6%) | 0.184 |
| Goggles use | 129/716; 18.0% | 65 (16.3%) | 64 (20.1%) | 0.189 |
| Shin Guard use | 75/716; 10.5% | 46 (11.6%) | 29 (9.1%) | 0.290 |
| Shielding of primary radiation |  |  |  |  |
| Collimation use | 446/594; 75.1 | 246 (67.6%) | 200 (87%) | <0.001 |
| RS: Radiation Safety. Responses with always used and mostly used are clubbed as appropriate use of RS practices. Rarely, never and not available were clubbed as practice that needs to change. |  |  |  |  |

**Table. 3.**
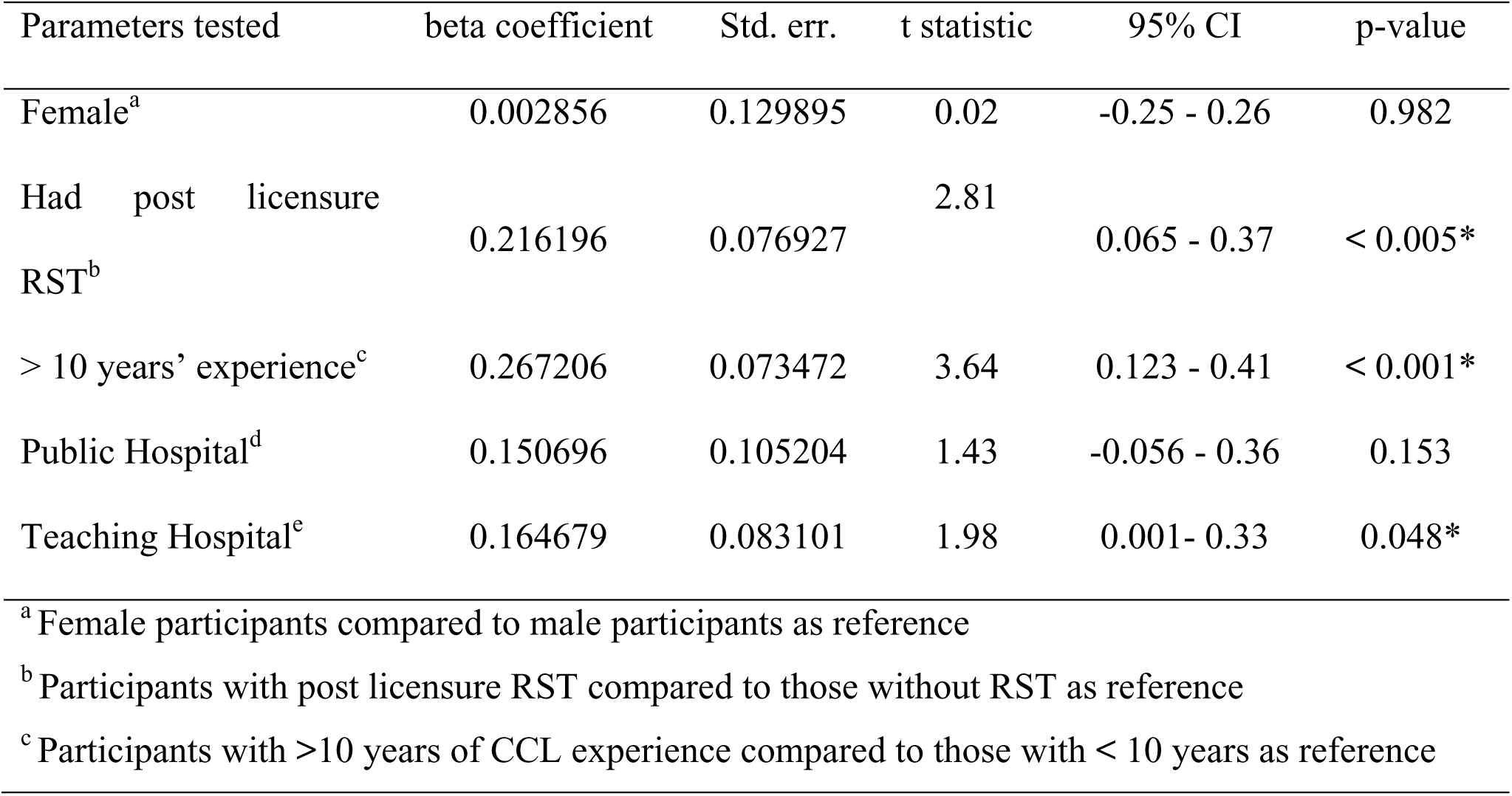
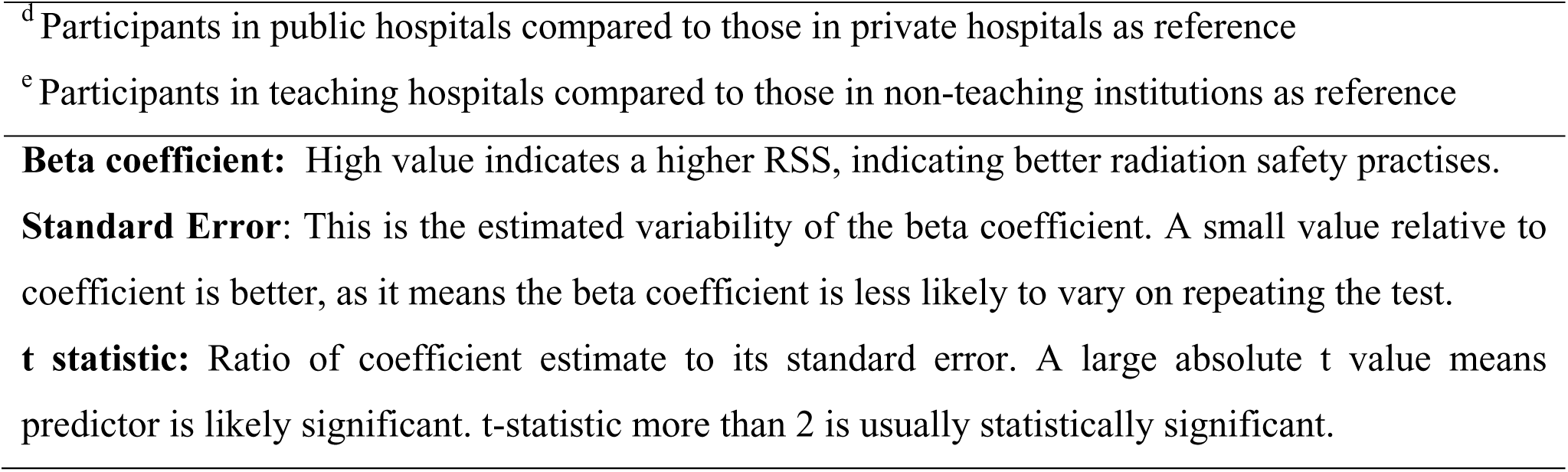
Multivariable log-linear regression model affecting the radiation safety score.

## Discussion

This nationwide, cross-sectional survey provides an overview of contemporary occupational radiation safety practices among interventional cardiologists in India, and highlights the critical importance of structured training programs in promoting protective behaviours, especially among early-career interventional practitioners. To our knowledge, this is one of the first studies to evaluate the impact of prior safety training programs held at the national level on the knowledge, attitudes, and practices of interventionalists. The key findings are: 1) current radiation safety practices remain suboptimal among interventional cardiologists in India and require significant improvement; 2) post-licensure radiation safety training is independently associated with higher radiation safety scores across all radiation safety domains tested. 3) greater years of professional experience was correlated with better adherence to essential radiation safety practices, though not necessarily with uniform use of all protective tools; and 4) practitioners in academic or teaching institutions demonstrated modestly better compliance with safety measures compared to those in non-teaching settings.

### Comparison with Existing Literature and Implications

International guidelines and expert consensus statements from various bodies, including the American Heart Association (AHA), International Commission on Radiological Protection (ICRP), Cardiovascular and Interventional Radiology Society of Europe (CIRSE), European Society for Vascular Surgery (ESVS), and the Society for Cardiovascular Angiography and Interventions (SCAI), provide clear, evidence-based recommendations for minimizing occupational exposure.^7–12^ Despite these well-established recommendations, our findings confirm the existence of a persistent “knowledge-to-practice gap” among interventional cardiologists in India. The mean RSS of 56.5 underscores the need for substantial improvement, and the fact that only 70% achieved moderate scores (≥50), and fewer than one in five scored above 75 reflects the true rarity of high proficiency.

Specific deficiencies were evident in key domains. Only 45% of participants reported using fluoroscopic frame rates ≤ 7.5 frames per second, despite robust evidence demonstrating up to 74% reductions in radiation exposure at these specific fluoroscopic settings.^13–15^ Likewise, although lead caps confer significant protection from cranial exposure even when ceiling-mounted shields are used,^16^ their adoption remained suboptimal (34%), with only a modest increase among those who had undergone post-licensure radiation safety training (36%).

Further, documentation of radiation dose in procedure reports was infrequent (23%), and one-fourth of respondents reported to never using collimation. These findings are concerning when considered against the backdrop of the PROTECTION VIII study which found that, although total patient radiation dose declined by 36% over a decade, operator radiation exposure remained high during complex PCI cases, even after accounting for technological advancements in radiation protection equipment.^17^ Combined with the results of our survey, these trends further underscore the urgent need for interventional cardiologists to prioritize personal radiation safety in the catheterization laboratory. Ensuring consistent adherence to protective measures is essential to safeguard both operators and patients, allowing both parties to benefit from safe procedural outcomes.

Orthopaedic morbidity related to heavy protective gear remains a significant occupational hazard. Indeed, nearly half of all respondents in our survey reported cervical or lumbar pain attributed to lead aprons. This rate is higher than that reported in European surveys, where 30.2% of interventional cardiologists cited musculoskeletal problems^3^ and nearly 20% of catheterisation laboratory professionals reported back, neck, or hip pain,^18^ potentially reflecting differences in workplace ergonomics, workload, and physical rehabilitation practices. The 2023 Occupational Health Hazards in Interventional Cardiology survey mirrored these grave findings and emphasized that orthopaedic injuries were highly prevalent at unacceptable levels (60%), a pattern that had changed little since the last survey conducted in 2014.^1^ Collectively, these findings emphasize the urgent need for ergonomic training,^19^ and the development of lighter protective gear. Cataracts and malignancies are known long-term adverse outcomes among interventionalists,^20,21^ but the former were infrequently reported in our cohort, likely reflecting under-recognition or limited surveillance. However, malignancies were not captured in our survey. Nonetheless, these findings highlight the importance of integrating routine dosimetry monitoring, occupational health review, and ophthalmologic screenings in safety training programs for healthcare professionals.

Our results are in concordance with previous Indian surveys reporting poor awareness and suboptimal adherence to safety practices^4,22^. Similar trends have been observed globally among interventional radiologists and cardiologists from Iran, Saudi Arabia, Portugal, Botswana, and the United Kingdom.^6,23–26^ Taken together, these studies reveal that inadequate knowledge and inconsistent use of radiation protection measures are universal challenges that transcend geography and resources.

Our study also demonstrated that greater professional experience was linked to better adherence to radiation safety practices. Clinicians with more than10 years of experience in the catheterization laboratory exhibited superior knowledge, greater training participation, and higher compliance with key safety measures, including personal dose monitoring, shield usage, and fluoroscopy time minimization. This trend likely reflects the cumulative professional exposure and evolving radiation safety awareness over time, rather than systematic incorporation of radiation safety principles during early training.

A key and novel finding of our study is the independent association between post-licensure radiation safety training and improved radiation safety performance across all domains.

Attending a radiation safety lecture/ training was associated with a 21.5% improvement in safety scores, which reflects an effect magnitude equivalent to the cumulative gain that would be accrued from 16 years of experience in the catheterization laboratory (1.29% increase in RSS per year of professional experience). This finding illustrates that structured training provides a rapid, tangible benefit that cannot be matched by experiential learning alone, which develops gradually over many years. Relying solely on the acquisition of safety habits during this timeframe is inadequate given that contemporary interventional cardiology is burdened with increasingly complex coronary and structural procedures that have long operating hours, which renders the interventional cardiologists prone to developing more radiation hazards over their 3-4 decades of practice. Therefore, early targeted training is essential to address persistent deficiencies, even among trained individuals, and institutions must necessitate periodic refresher courses, hands-on workshops, and simulation-based exercises. At present, however, there are no formal mandates from the National Medical Commission (NMC) or the Cardiological Society of India (CSI) that specify minimum training requirements for residents or consultants. On the other hand, nearly all

U.S. states require formal site-specific radiation safety training prior to procedural exposure.^28^ This contrast highlights an urgent opportunity for Indian professional societies and licensing authorities to implement standardized, mandatory curricula and certification programs in radiation safety. In light of these findings, we propose a concise, practical checklist named “The Ten Commandments of Radiation Safety” to guide implementation and audit of best practices across catheterization laboratories nationwide. Establishing such a framework is essential to foster a sustained culture of radiation safety and ensure long-term occupational health of all professionals.

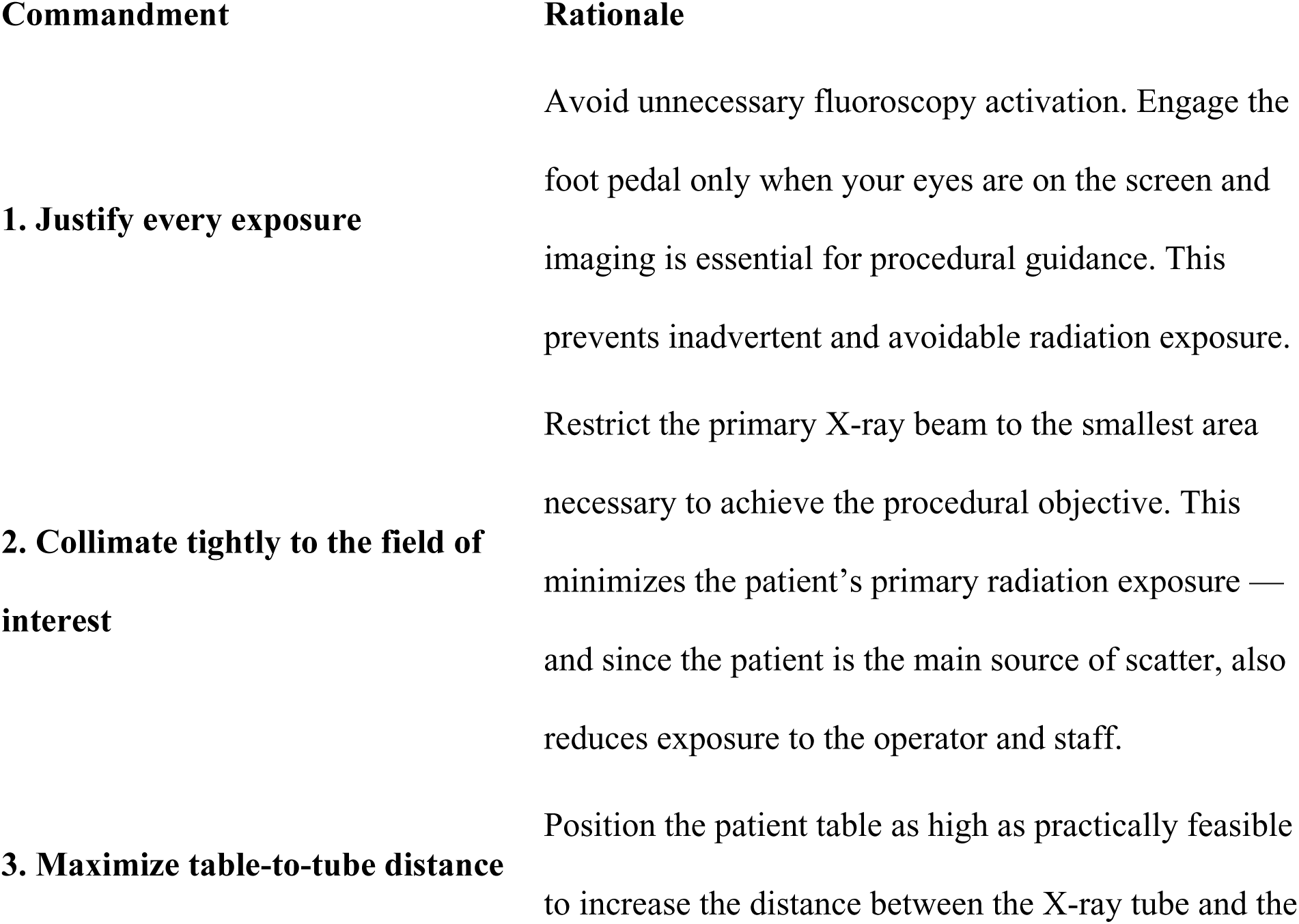
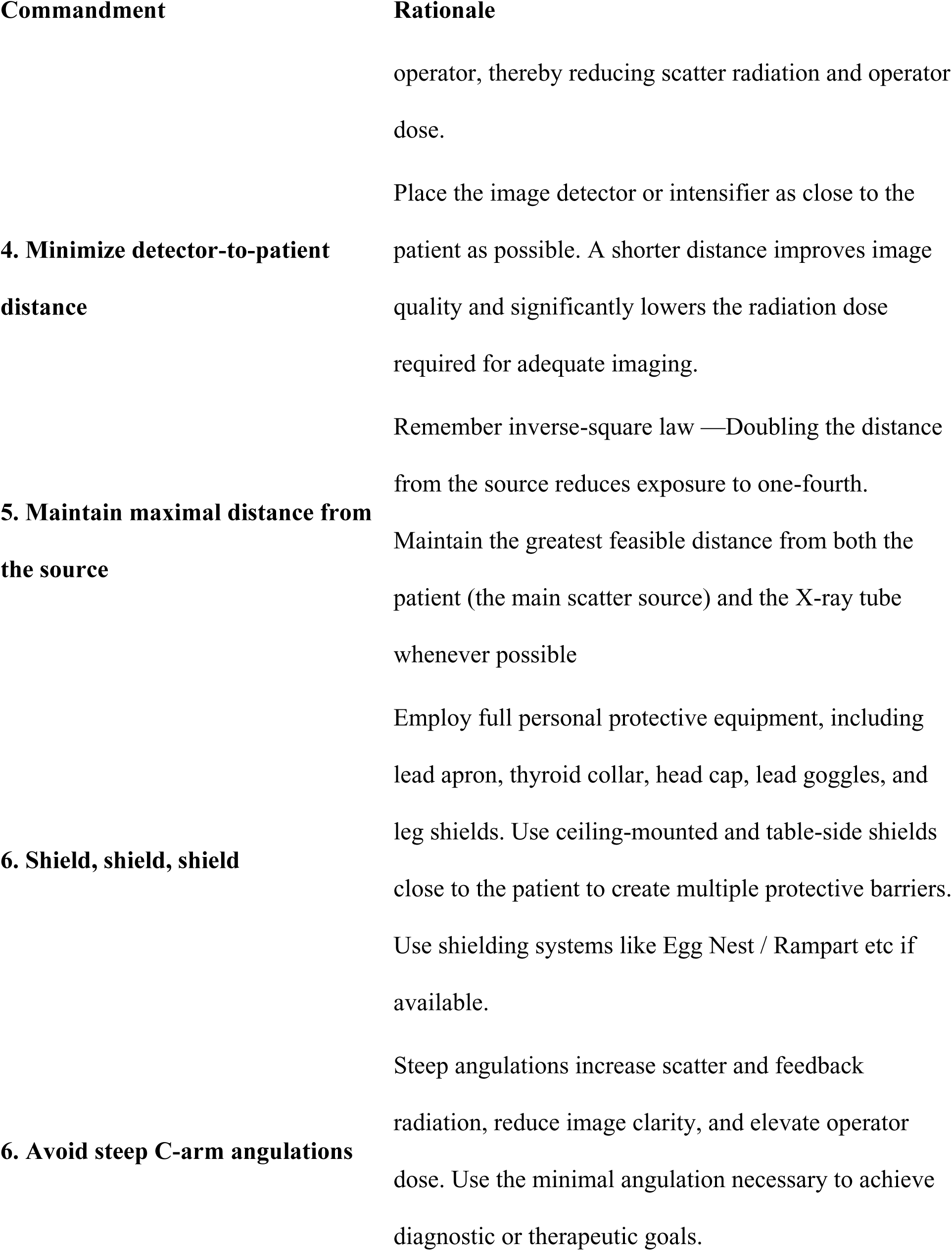
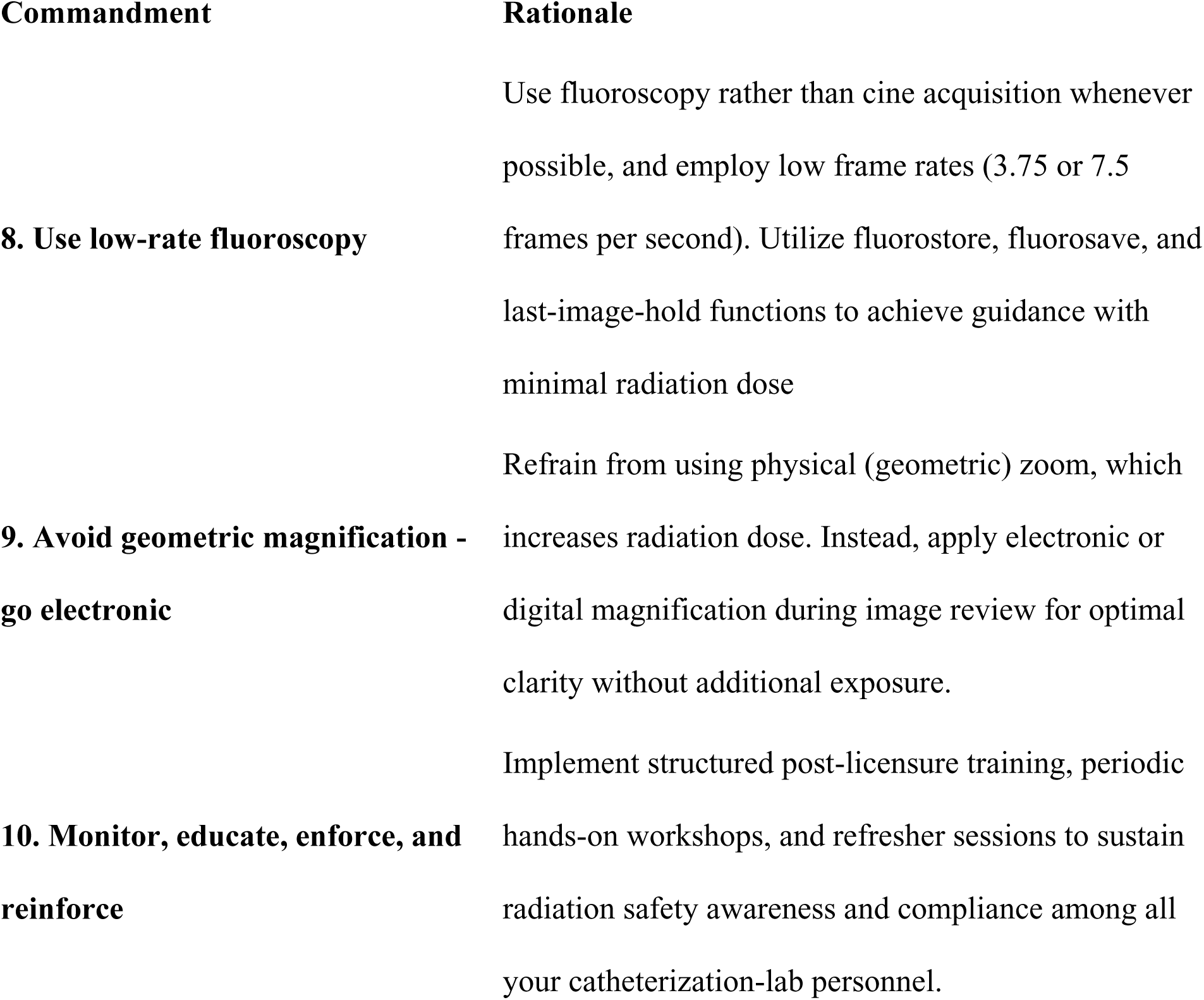

Engage the foot pedal only when your eyes are on the screen and imaging is essential for procedural guidance.

### Strengths, limitations, and future directions

This nationwide cross-sectional survey provides one of the first comprehensive assessments of occupational radiation safety knowledge, attitudes, and practices among interventional cardiologists in India. The broad geographic representation and inclusion of both academic and non-academic practitioners provide a credible overview of current real-world practices. The use of a structured, questionnaire enabled systematic evaluation of key domains, including knowledge, protective behaviors, and prior training exposure. However, several limitations must be acknowledged. First, as with all cross-sectional designs, causal inference cannot be established in our study. Although post-licensure radiation-safety training was strongly associated with higher radiation safety scores, we cannot definitively conclude that training unequivocally caused the observed improvements. Second, as a self-reported survey, responses are subject to recall, response, and social desirability biases, potentially leading to an overestimation of adherence to best practices. Third, the survey was conducted over different time points and meeting venues, with a modest response rate, which introduces the possibility of sampling bias and overrepresentation of more motivated or academically engaged participants. Fourth, the findings, while representative of India, may not be generalizable to high-income countries with established mandatory post-licensure radiation safety certification programs, though they are likely applicable to other low- and middle-income nations with similar training and practice environments. And finally, the Radiation Safety Score (RSS) was a composite index developed for this study and has not been externally validated against objective measures such as dosimetry data or patient/operator outcomes. The absence of longitudinal data precludes linking reported practices with actual radiation exposure or health outcomes. Future research should prioritize prospective, longitudinal studies aimed at linking radiation safety behaviours with actual occupational radiation doses and associated health outcomes in interventional cardiologists and supporting staff. Evaluating and incorporating interactive training models such as virtual reality simulations, active dosimetry feedback, and periodic refresher courses may help increase the uptake of competency-based certification. National-level registries and coordinated efforts by professional societies, regulatory agencies, and healthcare institutions are vital to foster a sustainable culture of radiation safety.

## Conclusion

This large, nationwide survey reveals substantial gaps in radiation safety practices among interventional cardiologists in India. The findings emphasize that structured post-licensing radiation safety training produces rapid, significant improvements in protective behaviours and may exceed gains achieved by experience alone. The magnitude of the training effect, though modest, argues strongly for institutionalizing radiation safety training as a recurrent, competency-based requirement rather than an optional or one-time educational event. To this end, regulatory bodies like the NMC should define formal minimum standards for radiation safety education, certification, and continuous professional development.

## Data Availability

The data underlying this study may be requested from the corresponding author. Fully de-identified data will be shared upon reasonable request and with approval from the Institutional Ethics Committee

## Acknowledgments

The authors would like to thank the organising committees of the various cardiology meeting including but not limited to Annual conference of Cardiological Society of India 2022, Tamil Nādu interventional council meeting 2023, National Intervention council meeting 2023.

